# Rapid Microscopic Fractional Anisotropy Imaging via an Optimized Kurtosis Formulation

**DOI:** 10.1101/2020.11.23.20237099

**Authors:** N. J. J. Arezza, D. H. Y. Tse, C. A. Baron

## Abstract

Water diffusion anisotropy in the human brain is affected by disease, trauma, and development. Microscopic fractional anisotropy (μFA) is a diffusion MRI (dMRI) metric that can quantify water diffusion anisotropy independent of neuron fiber orientation dispersion. However, there are several different techniques to estimate μFA and few have demonstrated full brain imaging capabilities within clinically viable scan times and resolutions. Here, we present an optimized spherical tensor encoding (STE) technique to acquire μFA directly from the 2nd order cumulant expansion of the dMRI signal (i.e. diffusion kurtosis) which requires fewer powder-averaged signals than other STE fitting techniques and can be rapidly computed. We found that the optimal dMRI parameters for white matter μFA imaging were a maximum b-value of 2000 s/mm^2^ and a ratio of isotropic to linear tensor encoded acquisitions of 1.7 for our system specifications. We then compared two implementations of the direct approach to the well-established gamma model in 4 healthy volunteers on a 3 Tesla system. One implementation of the direct cumulant approach used mean diffusivity (D) obtained from a 2nd order fit of the cumulant expansion, while the other used a linear estimation of D from the low b-values. Both implementations of the direct approach showed strong linear correlations with the gamma model (ρ=0.97 and ρ=0.90) but mean biases of −0.11 and −0.02 relative to the gamma model were also observed, respectively. All three μFA measurements showed good test-retest reliability (ρ≥0.79 and bias=0). To demonstrate the potential scan time advantage of the direct approach, 2 mm isotropic resolution μFA was demonstrated over a 10 cm slab using a subsampled data set with fewer powder-averaged signals that would correspond to a 3.3-minute scan. Accordingly, our results introduce an optimization procedure that has enabled clinically relevant, nearly full brain μFA in only several minutes.

**Highlights:**

- Demonstrated method to acquire optimal parameters for regression μFA imaging
- μFA measured using an optimized linear regression method at 3T
- First μFA comparison between direct regression approach and the gamma model
- Both approaches correlated strongly in white matter in healthy volunteers
- Nearly full brain μFA demonstrated in a 3.3-minute scan at 2 mm isotropic resolution

## 1. Introduction

Diffusion MRI (dMRI) can noninvasively acquire information about the microstructural characteristics of biological systems by probing the displacement of water molecules in tissue (Stejskal and Tanner, 1965; Tanner, 1965). Microstructural features that affect the apparent diffusion rate of water include cell size, shape, density, orientation, and the presence of membranes and barriers; thus, dMRI has found use in the study of neurological diseases that alter tissue microstructure (Inglese and Bester, 2010; Rovaris et al., 2005; van Everdingen et al., 1998; Zhang et al., 2009).

The most commonly used dMRI technique is diffusion tensor imaging (DTI) (Basser et al., 1994), in which dMRI data is fitted to the diffusion tensor model to estimate metrics such as the mean diffusivity (D) and fractional anisotropy (FA). DTI represents the dMRI signal as being entirely characterized by Gaussian diffusion (Basser, 1995), implicitly meaning the logarithm of the dMRI signal is assumed to depend on the b-value up to the first order in the cumulant expansion (Frisken, 2001). Despite DTI’s use in clinical applications, diffusion in tissues is too complex to be fully represented by Gaussian diffusion at high b-values (Johansen-Berg and Behrens, 2013), and characterizing the non-Gaussian effects provides more information about the underlying tissue (Shemesh et al., 2011). Diffusion kurtosis imaging (DKI) was developed to capture the effects of non-Gaussian diffusion by expanding the dMRI signal using cumulants up to second order in b-value (Jensen et al., 2005). Generally, DKI has been shown to be more sensitive than DTI towards quantifying microstructural changes that result from disease (Falangola et al., 2008; Fieremans et al., 2013; Wang et al., 2011).

Non-Gaussian diffusion can be attributed to a number of sources, such as polydisperse diffusion tensors with different mean diffusivities and diffusion tensors dispersed among multiple orientations (Henriques et al., 2020). Unfortunately, both DTI and DKI are unable to distinguish between true microstructural changes and neuron fiber orientation dispersion, reducing their specificity to disease in brain regions containing crossing or fanning axons (Jones et al., 2013; Szczepankiewicz et al., 2015). While DTI does not consider the effects of kurtosis at all, DKI cannot differentiate between the different sources of kurtosis without imposing assumptions about the underlying tissue (Fieremans et al., 2011; Jensen et al., 2005).

Of the many dMRI metrics, water diffusion anisotropy is particularly useful for studying neurological diseases because of the eccentric shape of neuronal axons. Despite confounding orientation dispersion with true microstructural characteristics in brain regions containing crossing or fanning neuron fibers, FA has been shown effective for detecting evidence of neurodegeneration in numerous diseases including multiple sclerosis (MS) (Ceccarelli et al., 2007; Cercignani et al., 2001), Alzheimer’s disease (Oishi et al., 2011), and stroke (Alegiani et al., 2017), among others. Generally, disease progression correlates with decreased anisotropy in white matter (WM) as neuronal axons lose their structural integrity or are demyelinated.

In recent years, efforts have been made to develop dMRI techniques that can quantify water diffusion anisotropy independent of neuron fiber orientation dispersion (Jespersen et al., 2013; Lasič et al., 2014). Microscopic anisotropy (μA) is an anisotropy metric that is independent of both reference frame and orientation dispersion, and microscopic fractional anisotropy (μFA) is a normalized variation of μA that additionally aims to remove the dependence on compartment size (Shemesh et al., 2016). There are multiple techniques to compute μFA, which can be categorized into: (1) methods that involve the use of linear tensor encoding (LTE) sequences (Kaden et al., 2016b, 2016a; Novikov et al., 2019), (2) methods that utilize double diffusion encoding (DDE) (Cory et al., 1990), and (3) methods that use nonconventional continuous gradient waveforms such as spherical tensor encoding (STE) (Eriksson et al., 2015; Lampinen et al., 2017; Lasič et al., 2014; Szczepankiewicz et al., 2015; Westin et al., 2016).

LTE methods utilize models to decouple microstructural properties from mesoscopic tissue orientation (Henriques et al., 2019). These techniques require prior knowledge or estimates of tissue properties such as the axonal volume fraction or the intracellular radial diffusivity (Henriques et al., 2019) but are highly accessible because LTE sequences are commonly used in both DTI and DKI. Generally, anisotropy can be estimated by acquiring LTE signals across multiple directions and b-shells and fitting the powder-averaged signals to a constrained model such as the spherical mean technique (SMT) model (Kaden et al., 2016b, 2016a). Recently, Henriques et al showed that μFA estimations using LTE are inaccurate compared to ground truth anisotropy, suggesting the techniques are not robust or do not sufficiently describe the underlying microstructure (Henriques et al., 2019).

DDE techniques to estimate μA and μFA use two independent diffusion-encoding pulse vectors in succession to probe the correlation of water diffusion in different directions (Ianuş et al., 2018; Jespersen et al., 2013; Lawrenz and Finsterbusch, 2015; Mitra, 1995; Ozarslan and Basser, 2008). DDE can distinguish between microstructural properties and orientation dispersion without imposing modeling constraints (Cory et al., 1990; Mitra, 1995), likely making the technique more robust and accurate than LTE techniques by eliminating the possibility of assumption misestimation. Furthermore, the clinical viability of DDE μFA imaging was demonstrated in a preliminary study of MS patients at 3T with a 5 minute scan time and 3 mm isotropic resolution (Yang et al., 2018). While DDE is a promising technique, it has some limitations. Due to the use of two consecutive diffusion-encoding pulses separated by a mixing time, DDE sequences require longer TEs than standard LTE sequences to achieve equal b-values. Furthermore, a twice-refocused implementation is required to avoid biases due to concomitant fields (Baron et al., 2012; Szczepankiewicz et al., 2019), further increasing the TE.

Techniques that utilize nonconventional diffusion-encoding waveforms probe unique q-space trajectories that provide additional information about tissue microstructure beyond the capabilities of LTE. One technique to estimate μFA and other parameters is the gamma model, in which the inverse Laplace transform of the gamma distribution is fitted to powder averaged dMRI signals from LTE acquisitions and STE acquisitions (Szczepankiewicz et al., 2016, 2015). In the gamma model, signal variance due to non-Gaussian diffusion is characterized into two sources: isotropic variance arising from polydispersity in mean diffusivity, and anisotropic variance arising from microscopic anisotropy (Szczepankiewicz et al., 2015); the model assumes that LTE signal depends on both isotropic and anisotropic variance while STE signals depend only on isotropic variance. Gamma model μFA protocols use unique waveforms to acquire single-shot isotropic diffusion weighted signals (Eriksson et al., 2013; Lasič et al., 2014). Though STE is more TE-efficient than DDE, STE waveforms can potentially introduce time-dependent effects due to varying spectral content over the different gradient channels (Eriksson et al., 2013). Furthermore, the implementation of the gamma model described by Szczepankiewicz et al (Szczepankiewicz et al., 2015) is a computationally-intensive technique for parameter estimation. Other techniques include deriving μFA from higher order cumulant expansions with direct linear regression of the cumulant expansion of the diffusion signal (Nery et al., 2019) and the correlation tensor model (Henriques et al., 2020). Notably, the direct linear regression approach offers the potential advantages of computational efficiency over the gamma model, but the two have not been directly compared.

The application of μFA imaging to clinical workflow is appealing due to the unique insight it provides into brain microstructure. The parameter’s insensitivity to neuron fiber orientation is advantageous over FA in the study or diagnosis of neuropathology in brain regions containing crossing or fanning fibers. However, μFA generally requires long scan times that are not clinically feasible, especially when used in conjunction with other imaging techniques that are required in the clinical workflow. Other demonstrations of μFA that have achieved shorter scan times did so at the cost of resolution (Nilsson et al., 2019; Yang et al., 2018), producing μFA maps with poorer resolution than typical FA maps acquired with DTI. In order to maximize scan efficiency, it is essential to understand the optimal parameters required to measure μFA. To our knowledge, no comprehensive assessment of the optimal choices of b-value and relative numbers of LTE and STE acquisitions for a direct linear regression approach have been performed.

In this work, we investigated the optimal b-values and ratio of STE to LTE acquisitions for the estimation of μFA in white matter. We combined these findings with two implementations of direct linear regression to enable the acquisition of full-brain, 2 mm isotropic resolution μA and μFA maps *in vivo* within a 4.5 min scan time and a 2-minute computation time. Estimates of μFA using direct approaches strongly correlated with the gamma model (ρ ≥ 0.9), and all approaches exhibited high test-retest reliability (ρ ≥ 0.77).

## 2. Theory

### 2.1 μFA estimation

The normalized signal intensity of powder-averaged dMRI acquisitions can be represented by the cumulant expansion (Lasič et al., 2014):

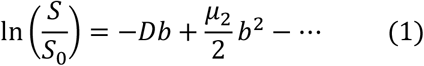

where *S* is the powder-averaged signal, *S*_*0*_ is the mean signal with no diffusion encoding, *b* is the b-value, and *μ*_*2*_ is the second central moment or variance. Lasic et al (Lasič et al., 2014) define the microscopic fractional anisotropy in terms of the scaled difference in variance between powder-average LTE and STE acquisitions:

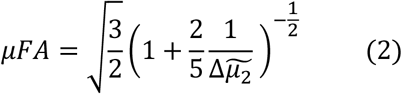

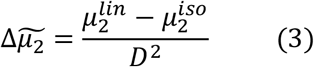

where 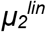 and 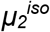 are the second terms in the cumulant expansions of powder-averaged LTE and STE acquisitions, respectively. Using equation (1) up to the second cumulant term, the powder-averaged linear and mean isotropic signals can be represented as:

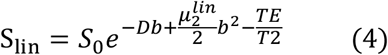

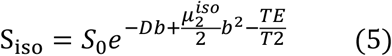

If it is assumed that the only sources of kurtosis are dispersion in pore size and orientation, then the diffusion coefficient D will be equal between LTE and STE (Szczepankiewicz et al., 2015). By assuming D is the same between LTE and STE signals acquired at the same b-value, Equations (4) and (5) can be substituted into equation (3) to provide an estimate of the scaled difference in variance that notably does not depend on the non-diffusion weighted signal *S*_*0*_:

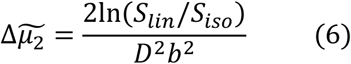

Substituting equation (6) into equation (1) provides an estimate of the μFA (Nery et al., 2019):

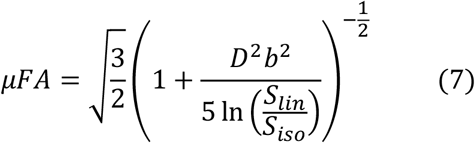

Microscopic anisotropy is defined here based on the difference in signal between linear and isotropic dMRI acquisitions, similar to the equation used in DDE protocols (Ianuş et al., 2018):

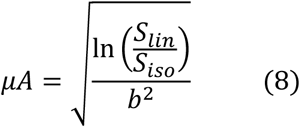

By ignoring the third and higher order cumulant terms in deriving equations (4) and (5), μA can be estimated from a single b-shell, reducing scan time; however, ignoring the higher cumulants comes with the cost of potentially introducing a bias to the measurement (Shemesh, 2018). μFA can then be expressed in terms of μA by substituting equation (8) into equation (7):

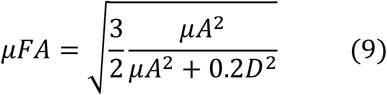

### 2.2 Diffusion coefficient estimation using the diffusion kurtosis model

Explicitly enforcing that the diffusion coefficient D is the same between LTE and STE acquisitions causes the minimum number of powder-averaged samples required to estimate D, 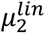 and 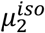 in a joint least squares estimation to be only 4 (with at least one non-zero b-value sampled for each of LTE and STE). Accordingly, a protocol to map μFA using equation (9) could contain LTE and STE acquisitions at a single high b-value for computation of μA^2^ using equation (8), plus either STE or LTE acquisitions at two smaller b-values to enable estimation of D, 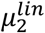, and 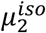 in a joint kurtosis model. It is also possible to estimate D from a linear fit over the low b-values, but this may introduce a bias. Notably, it is also possible to estimate uA^2^ directly from 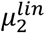 and 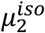 (equation (3)).

### 2.3 μA Optimization

To optimize a protocol for μA and μFA, sequence parameters that maximize the ratio of the mean measurement to its standard deviation can be evaluated, similar to the approach used to determine optimal parameters for diffusivity measurements (Xing et al., 1997). Using standard error propagation (Bevington et al., 1993), the signal-to-noise ratio (SNR) of a μFA image generated using equation (9) can be expressed as:

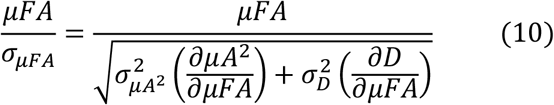

μFA image quality thus increases with reduced variance in μA^2^ and D measurements. It is expected that μA^2^ will generally have much higher variance than D because it depends only on the highest b-shell data (equation (8)), which has the lowest SNR. Thus, we will focus on the optimization of μA^2^ as a surrogate for the optimization of μFA. The SNR of a μA^2^ image can be expressed as (Appendix):

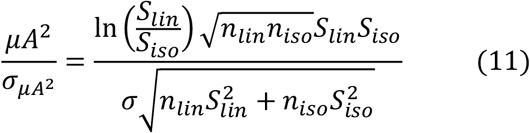

where *n*_*lin*_ is the number of LTE directions acquired, *n*_*iso*_ is the number of STE averages acquired, *S*_*lin*_ and *S*_*iso*_ are the powder-averaged signals of the LTE and STE images, respectively, and *σ* is the mean image noise. Given that *μA*^*2*^*/σ*_*μA2*_ is maximized when *n*_*iso*_/*n*_*lin*_ = *S*_*lin*_*/S*_*iso*_ (see Appendix), and that *S*_*lin*_ and *S*_*iso*_ are dependent on b-value, the optimal protocol parameters (*b* and *n*_*iso*_/*n*_*lin*_) can be determined using equation (11).

To observe the effect of TE on the μA^2^ SNR, equations (4) and (5) can be substituted into equation (11):

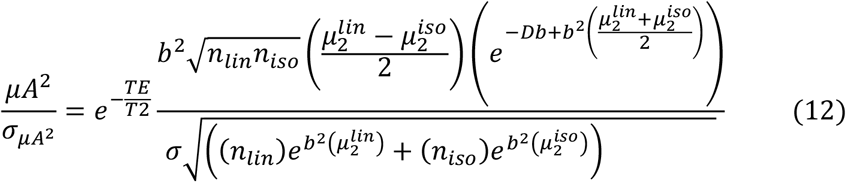

Equation (12) reveals that the SNR depends on TE by an exponential prefactor.

## 3. Methods

Two sets of MRI scans were performed on two sets of volunteers for this study. The first set of scans (3.1) consisted of LTE and STE acquisitions over a wide range of b-values and was acquired to provide the signal data needed to optimize μA using equation (11). The second set of scans (3.2, 3.3) performed test-retest measurements with a comprehensive sequence that allowed for μFA mapping using the gamma model, equation (2), and equation (9).

### 3.1 Sequence optimization

MRI scans were performed in 4 healthy volunteers (2 females, mean age 22.4 ± 1.7 years) on a 3T Prisma whole-body MR system (Siemens Healthineers) with 80 mT/m strength and 200 T/m/s slew rate. Informed consent was obtained before scanning and the study was approved by the Institutional Review Board at Western University. Multiple b-shell diffusion data were acquired in a single scan using LTE and STE sequences (Fig. 1): 6 image volumes were acquired at b = 0 s/mm^2^, and 6 linear directions and 6 isotropic averages were acquired at b-values between 500 and 3500 s/mm^2^, in increments of 500 s/mm^2^. The other parameters were TE/TR = 125/8700 ms, FOV = 192×192 mm^2^, 2 mm isotropic resolution, 45 slices, rate 2 GRAPPA, 2 averages, and total scan time = 29 minutes. Images were processed using Gibbs ringing correction and Eddy current correction with FSL Eddy (Andersson and Sotiropoulos, 2016).

**Fig. 1.**
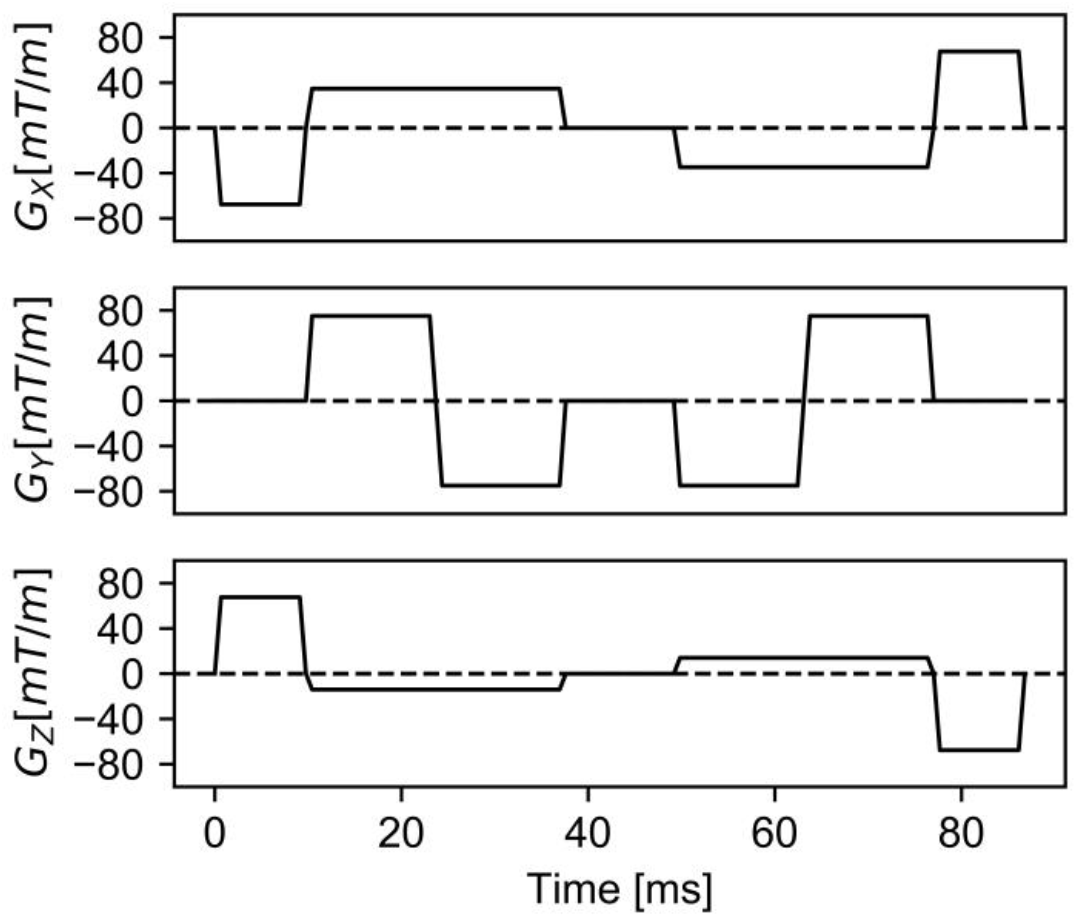
Schematic representation of the spherical tensor encoding gradient waveforms: diffusion encoding blocks have been inserted on both sides of a 180° pulse in all three gradient directions to acquire an isotropic diffusion MRI signal. Implicit gradient reversal due to the 180° pulse has been applied.

A region of interest (ROI) across multiple slices was manually selected in the frontal WM for each patient and used to measure the mean linear signal and mean isotropic signal at each b-value. A joint regression was performed on the mean LTE and STE signal data to fit the curves to equation (1) up to the third cumulant, with the assumption that D is the same in both linear and isotropic acquisitions. The best-fit cumulant expansions for each of the 4 volunteers were averaged and used together with equation (11) to determine the optimal b-value and optimal ratio of linear to isotropic acquisitions in a μA protocol. In evaluation of equation (11), the T2 decay constant was assumed to be 80 ms to approximate WM at 3T (Wansapura et al., 1999). These SNR calculations assume the same total number of acquisitions at each b-value, with only the ratio of *n*_*iso*_/*n*_*lin*_ acquisitions changing.

### 3.2 Test-retest Acquisitions

A comprehensive 113 acquisition dMRI protocol was used to acquire the data to compare μFA volumes generated with different methods. 4 healthy volunteers (2 females, mean age 28.0 ± 6.6 years) were imaged at 3T with a 9-minute dMRI scan with TE/TR = 94/4500 ms. The scan consisted of 3, 3, 15, 6, and 22 LTE directions and 6, 6, 10, 10, and 27 STE averages at b = 100, 700, 1000, 1400, and 2000 s/mm^2^, respectively, as well as 5 averages at b = 0 s/mm^2^. These directions were chosen to enable retrospective splitting of the data into the subsets described below. The other parameters were FOV = 220×220 mm^2^, 2 mm isotropic resolution, 48 slices, and rate 2 GRAPPA. Volunteers were also scanned using T1-weighted MPRAGE with 1 mm isotropic resolution. After removing each volunteer from the MR scanner for a period of 5-10 minutes, a repeat measurement was performed using only the dMRI protocol. Data from these acquisitions is available online [dataset] (Baron and Arezza, 2020).

Two separate post-processing pipelines were performed on the data to acquire two different data sets: a denoised data set to compare the direct μFA approaches to the gamma model, and a “noisy” data set that omitted denoising to test the effects of using an optimized vs. suboptimal ratio of STE to LTE scans to compute μA, since denoising is a non-linear operation that invalidates the assumptions used in the derivation of equation (11). All the diffusion MRI data was processed using Gibbs ringing correction and FSL Eddy <u>(Andersson and Sotiropoulos, 2016)</u>, and PCA denoising (Veraart et al., 2016) was performed prior to these corrections for the denoised data set.

The T1-weighted anatomical volumes were segmented into WM and gray matter (GM) masks using FMRIB’s Automated Segmentation Tool (FAST) (Zhang et al., 2001) and were registered to the denoised dMRI volumes using symmetric diffeomorphic and affine transforms with ANTS software (https://github.com/ANTsX/ANTs) (Chen et al., 2019). The retest noisy and denoise volumes were also registered to the respective test volumes using a rigid transform with ANTS.

The noisy dMRI data were split into two 56-acquisition subsets to represent optimized and suboptimal protocols, respectively, to validate equation (11). The optimized protocol was based on a rapid sequence proposed by Nilsson et al (Nilsson et al., 2019) and included 3, 3, 6, and 6 LTE directions and 6, 6, 10, and 16 STE averages at b = 100, 700, 1400, and 2000 s/mm^2^. The suboptimal protocol consisted of the same acquisitions with one exception: the ratio *n*_*iso*_/*n*_*lin*_ at the b = 2000 s/mm^2^ shell was 6/16 instead of 16/6, a suboptimal ratio (see 4.1). The 6 direction subset of LTE acquisitions used an icosahedral sampling scheme (Nilsson et al., 2019), and the 16 direction subset was distributed using electrostatic repulsion (Jones et al., 1999).

The denoised dMRI data were split into two subsets with each containing 56 acquisitions. The “standard subset”, to be used to compare the gamma model vs. equation (3), used the rapid sequence by Nilsson et al described above (Nilsson et al., 2019). An additional subset, referred to herein as the “DTI subset”, included 22 STE directions at b = 2000 s/mm^2^ and 3, 15, and 16 LTE averages at b = 100, 1000, and 2000 s/mm^2^ (56 total acquisitions), and was designed to investigate whether a single b-shell to compute μA^2^ (b = 2000 s/mm^2^) can be added to a DTI acquisition (b = 100, 1000 s/mm^2^) to enable μFA imaging using equation (9). The 15 and 16 direction LTE shells were determined separately from each other using electrostatic repulsion.

### 3.3 Test-retest Analysis

To compare the SNR of μA^2^ between the optimized and suboptimal subsets of the noisy dMRI data, μA^2^ was estimated at b = 2000 s/mm^2^ in both the test and retest volumes for each volunteer. The test-retest coefficients of variance (CoVs) of the optimized and suboptimal volumes across all volunteers were compared.

For the denoised data, the powder-averaged STE and LTE signals vs. b-value were fitted to the diffusion kurtosis model using a joint non-negative least squares method assuming consistent D between STE and LTE, and μFA was computed using equation (2) (μFA_E2_). μFA was also estimated using Nilsson et al’s Multidimensional diffusion MRI software (Nilsson et al., 2018) (https://github.com/markus-nilsson/md-dmri) to fit the diffusion-weighted signals to the gamma model (μFA_gamma_). μFA maps were generated for each volunteer using these two methods in the standard subset of data.

Additionally, μFA was estimated using equation (9) in the DTI subset by decoupling μA^2^ and D (μFA_E9+DTI_): μA^2^ was estimated at b = 2000 s/mm^2^ using the direct cumulant method (equation (8)) while D was estimated by fitting the b = 100 and 1000 s/mm^2^ data to the DTI model using FMRIB’s DTIFIT tool.

The μFA maps from the different methods and subsets were then compared in WM using Bland-Altman plots and voxelwise scatter plots, and Pearson correlation coefficients were computed between each technique. To test the repeatability of the measurement techniques, Bland-Altman plots were generated for each patient to compare the initial and repeat μFA volumes and Pearson correlation coefficients were computed between initial and repeat μFA maps.

## 4. Results

### 4.1 Sequence optimization

The logarithm of the powder-averaged WM dMRI signal as a function of b-value, averaged across all volunteers, is shown in Fig. 2. As expected (Szczepankiewicz et al., 2015), the departure from monoexponential signal decay was greater in the LTE than STE signal curve due to the mesoscopic orientation of tensors. Fig. 3 shows the variation in *μA*^*2*^*/σ*_*μA2*_ with b-value and the ratio of *n*_*iso*_/*n*_*lin*_. For any given b-value, the optimal *n*_*iso*_/*n*_*lin*_ was computed to be equal to the ratio of the powder averaged signals, *S*_*lin*_/*S*_*iso*_, at said b-shell. The highest *μA*^*2*^*/σ*_*μA2*_ occurred when the b-value was 2000 s/mm^2^, for which the optimal *n*_*iso*_/*n*_*lin*_ was approximately 1.7. However, a wide range of dMRI parameter configurations yielded an SNR above 95% of the optimal parameters for *μA*^*2*^ SNR.

**Fig. 2.**
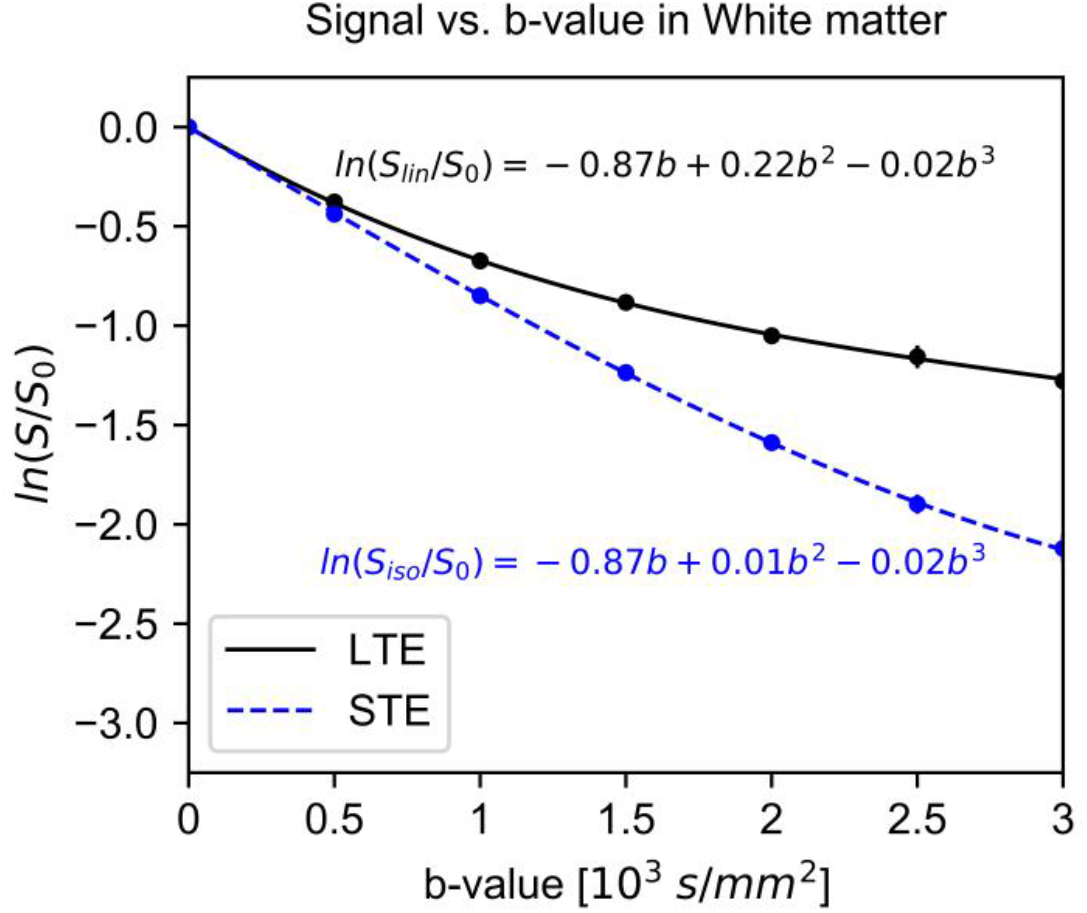
Logarithm of the diffusion MRI signal vs. b-value in frontal white matter. The plot shows the powder-averaged signal from a manually prescribed region of interest across four volunteers as measured with linear tensor encoding and spherical tensor encoding (black and blue circles, respectively), while the dashed lines show the third order cumulant model fit. Also depicted are the standard deviations across the volunteers.

**Fig. 3.**
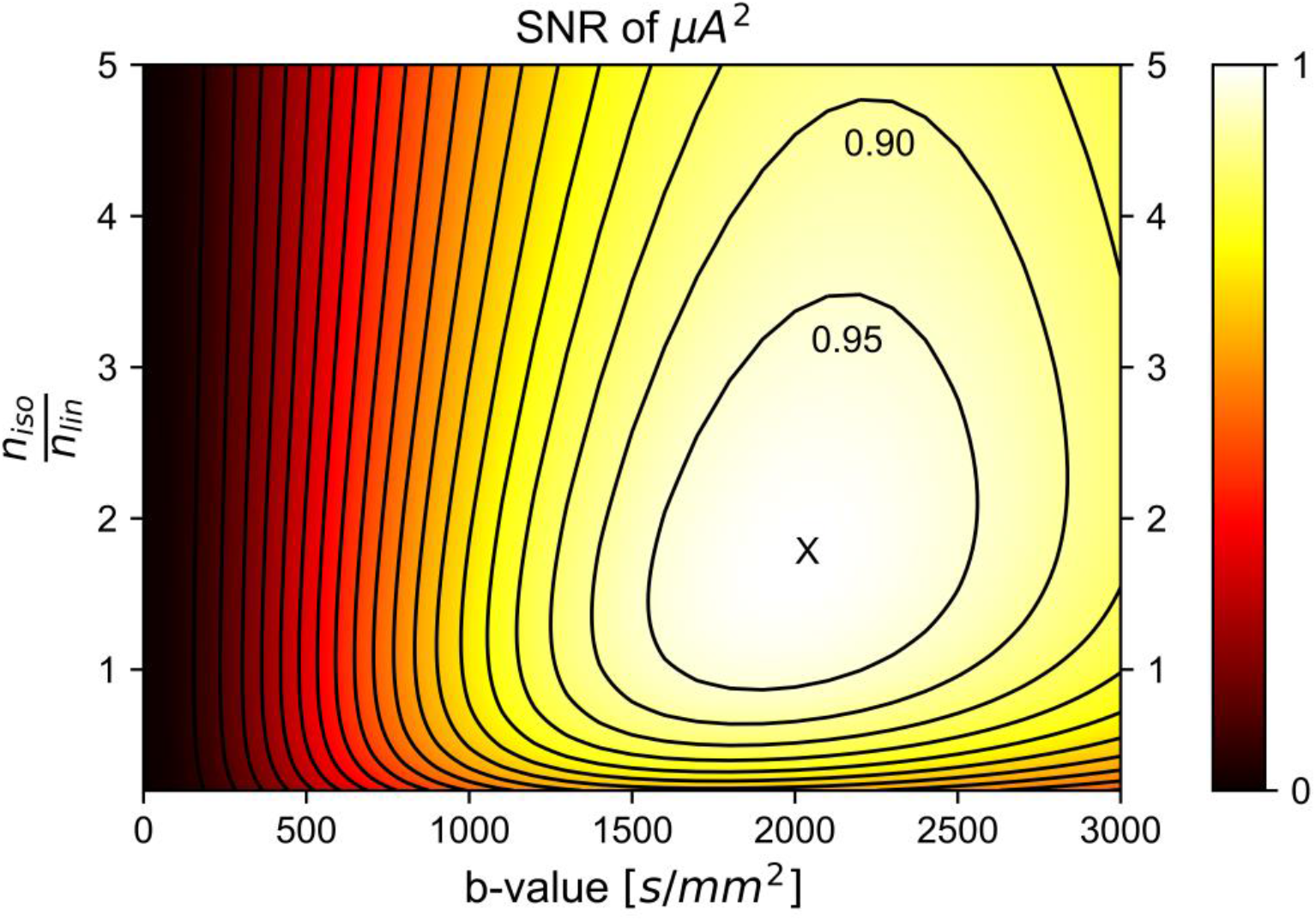
Simulated μA^2^ SNR in white matter as a function of the b-value and the ratio of STE to LTE acquisitions (*n*_*iso*_/*n*_*lin*_). Though the maximum SNR occurred when b = 2000 s/mm and *n*_*iso*_/*n*_*lin*_ = 1.7 (marked by an ‘X’), a wide range of parameters yielded SNRs greater than 95% of the maximum SNR, suggesting that there is flexibility in parameter choice when designing a protocol. Notably, a significant drop off in SNR occurred for *n*_*iso*_/*n*_*lin*_ < 1, suggesting that image quality is maximized when the number of STE acquisitions is greater than or equal to the number of LTE acquisitions.

Using the powder averaged STE and LTE WM signal data from the noisy data subset at b = 2000 s/mm^2^ across all volunteers along with equation (11), the SNR of μA^2^ in the suboptimal subset was predicted to be 87% of the SNR of μA^2^ in the optimized subset. Analysis of the test and retest μA^2^ volumes revealed a CoV of 22.96% in the optimized measurement and a CoV of 25.75% in the suboptimal measurement, yielding an experimentally acquired SNR ratio of approximately 89% (since CoV is analogous to SNR^-1^). Example μA^2^ images estimated using the optimized and suboptimal subsets are depicted in Fig. 4.

**Fig. 4.**
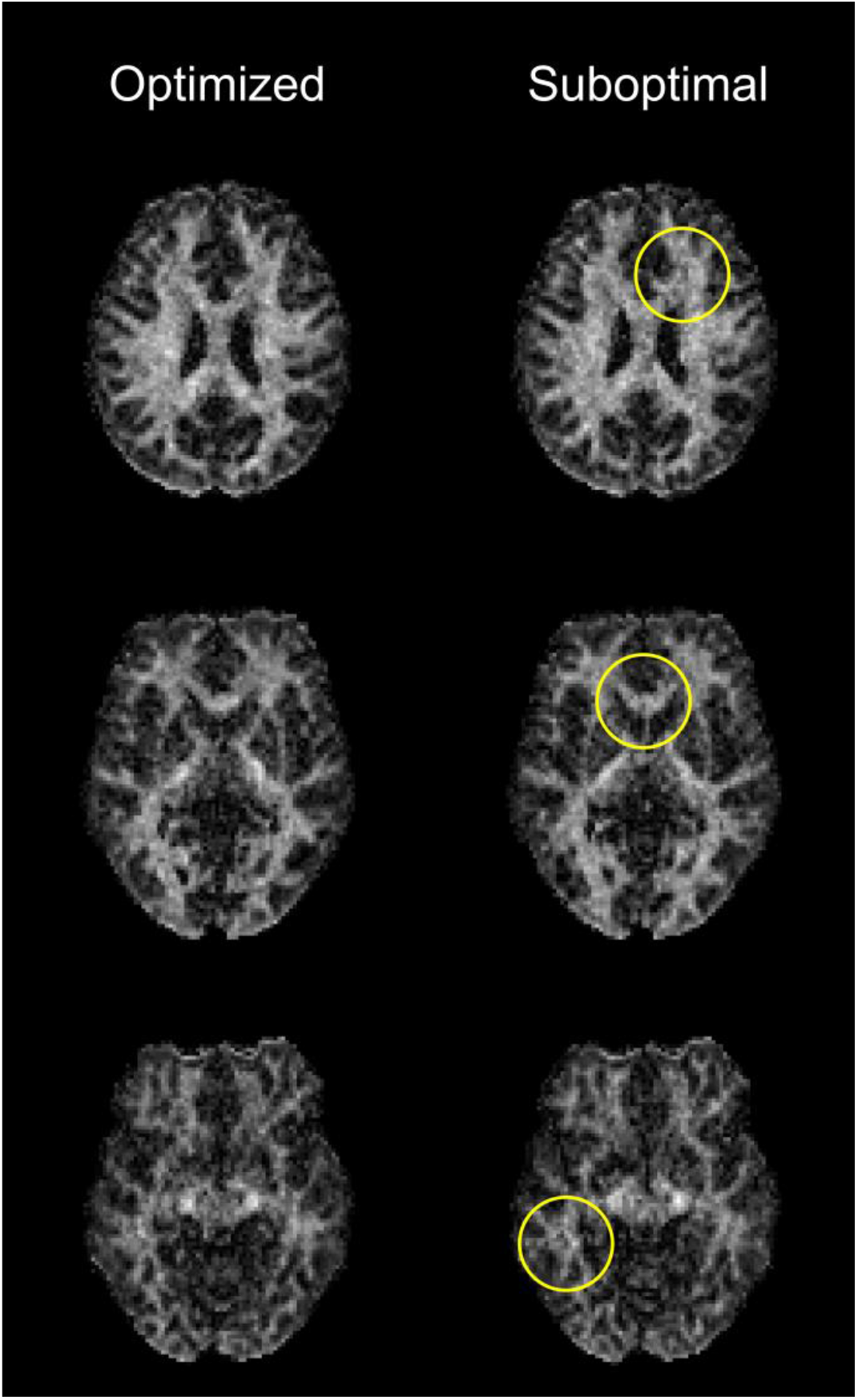
Example μA^2^ images acquired with the optimized (left) and suboptimal (right) subsets of the data without denoising. Lower image quality is observed in the right case, with some irregular features highlighted by the yellow circles.

### 4.2 Comparison between different μFA techniques

Example μFA_gamma_ and μFA_E2_ maps computed from the standard subset, as well as μFA_E9+DTI_ maps computed from the DTI subset, are depicted in Fig. 5. Aside from CSF regions, μFA is qualitatively consistent across the different techniques and data subsets and image quality is comparable between them.

**Fig. 5.**
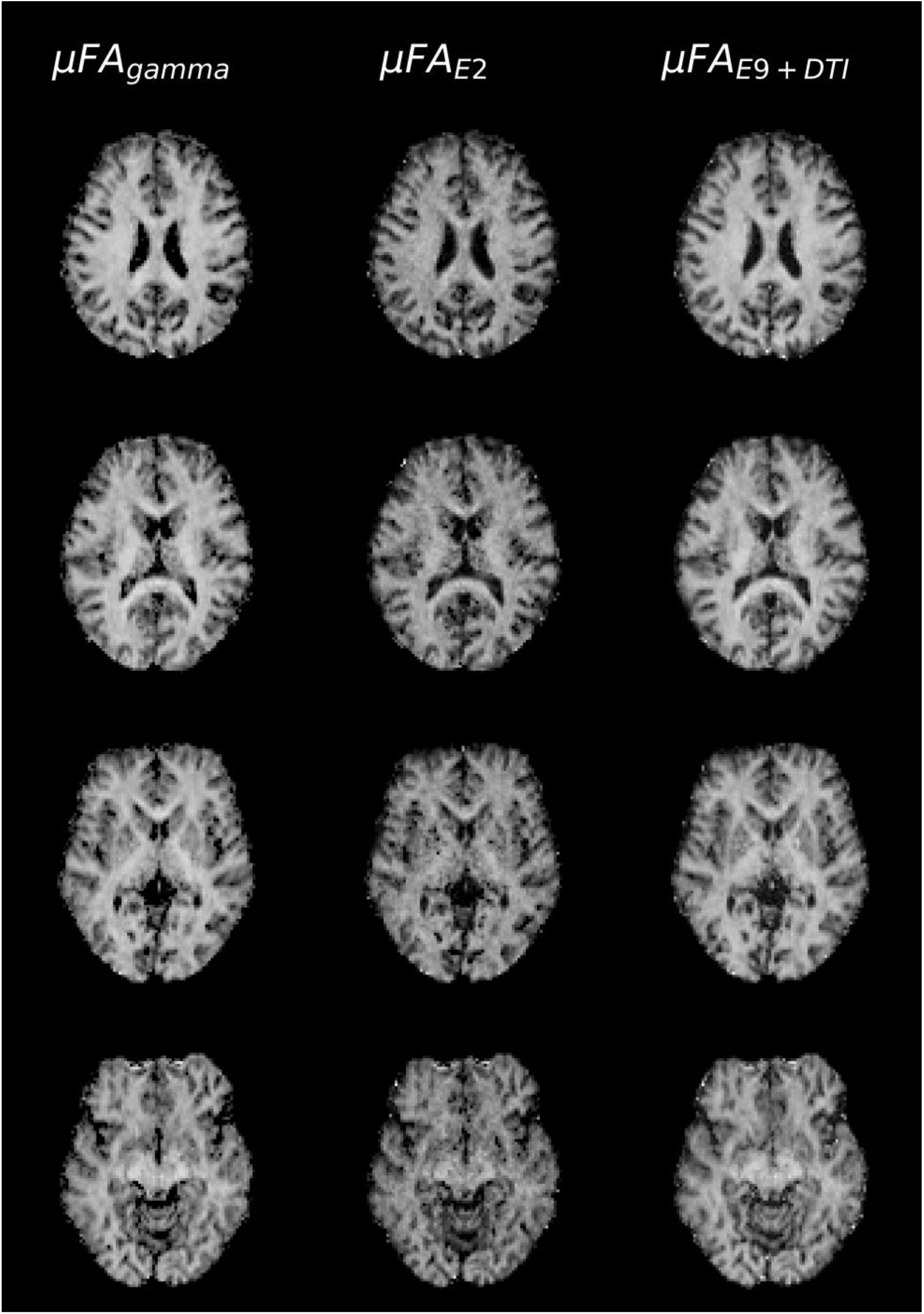
Example μFA images from one volunteer. Images were acquired using the gamma model with the standard subset (left), equation (2) with the standard subset (center), and equation (9) with MD computed from DTI using only b-values of 100 and 1000 s/mm^2^ (right). Comparable image quality is observed for the three methods.

Scatter plots and Bland-Altman plots comparing WM μFA using the three different estimation approaches in all volunteers are presented in Fig. 6. Strong linear correlations were observed in the scatter plots comparing each volume, with respective Pearson correlation coefficients of 0.97 (μFA_gamma_ vs. μFA_E2_), 0.90 (μFA_gamma_ vs. μFA_E9+DTI_), and 0.90 (μFA_E2_ vs. μFA_E9+DTI_). Relative to μFA_gamma_, the mean WM biases in the other volumes were −0.11 (μFA_E2_) and −0.02 (μFA_E9+DTI_).

**Fig. 6.**
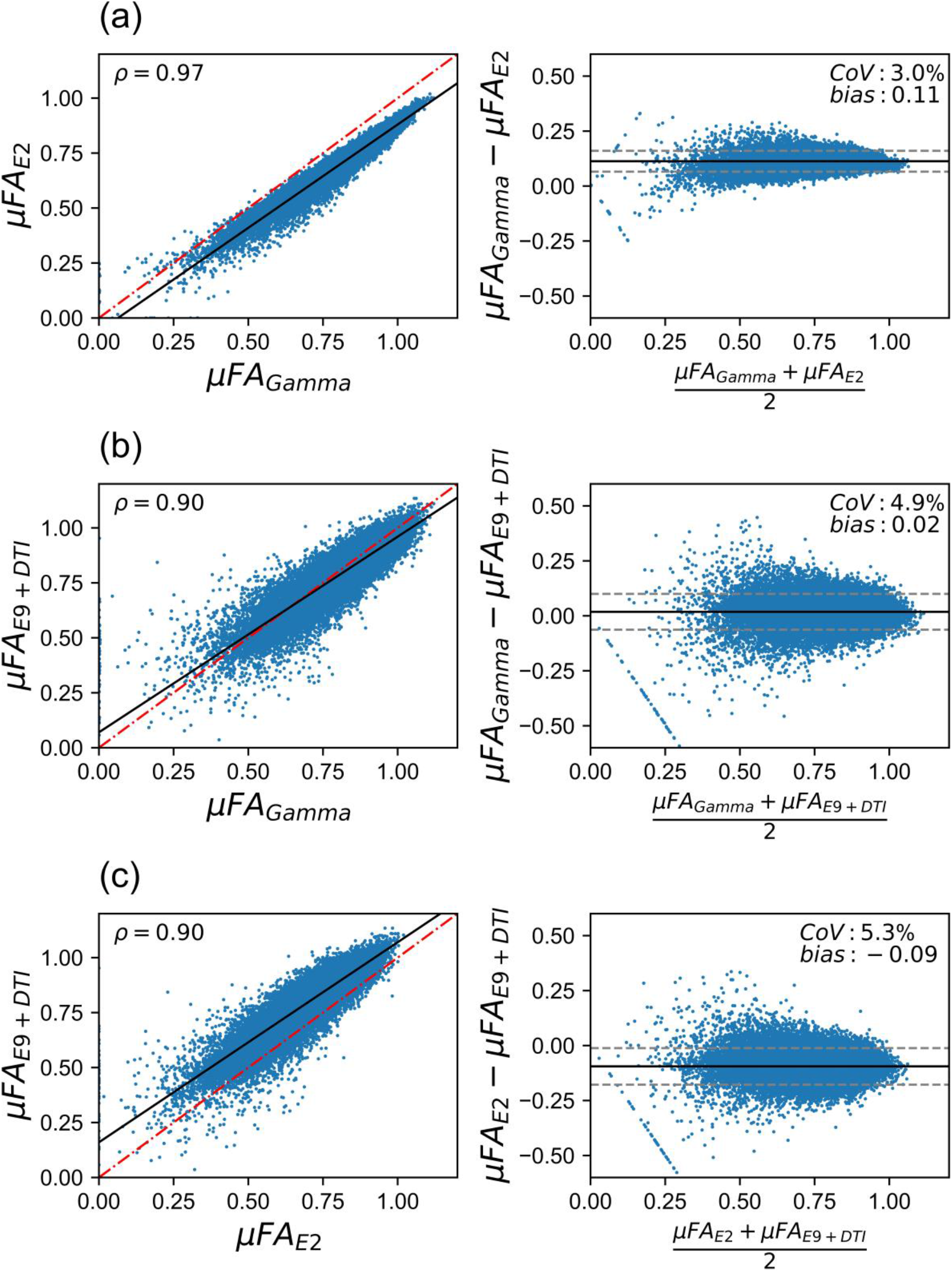
Voxelwise correlations between μFA estimates acquired using different techniques (left) and Bland-Altman plots depicting biases between the methods (right): (a) μFA_gamma_ vs. μFA_E2_, (b) μFA_gamma_ vs. μFA_E9+DTI_, and (c) μFA_E2_ vs. μFA_E9+DTI_. The dashed red line and solid black line in each of the scatter plots represent the identity and regression lines, respectively. The solid black line in the Bland-Altman plots represents the mean bias, and the dashed grey lines represent the ±1.96 standard deviation lines.

### 4.3 Analysis of repeatability

Bland-Altman plots comparing the test and retest μFA volumes across all volunteers revealed no biases in repeat measurements (Fig. 7). The Pearson correlation coefficients between the test and retest μFA maps were 0.83 (μFA_gamma_), 0.79 (μFA_E2_), and 0.84 (μFA_E9+DTI_).

**Fig. 7.**
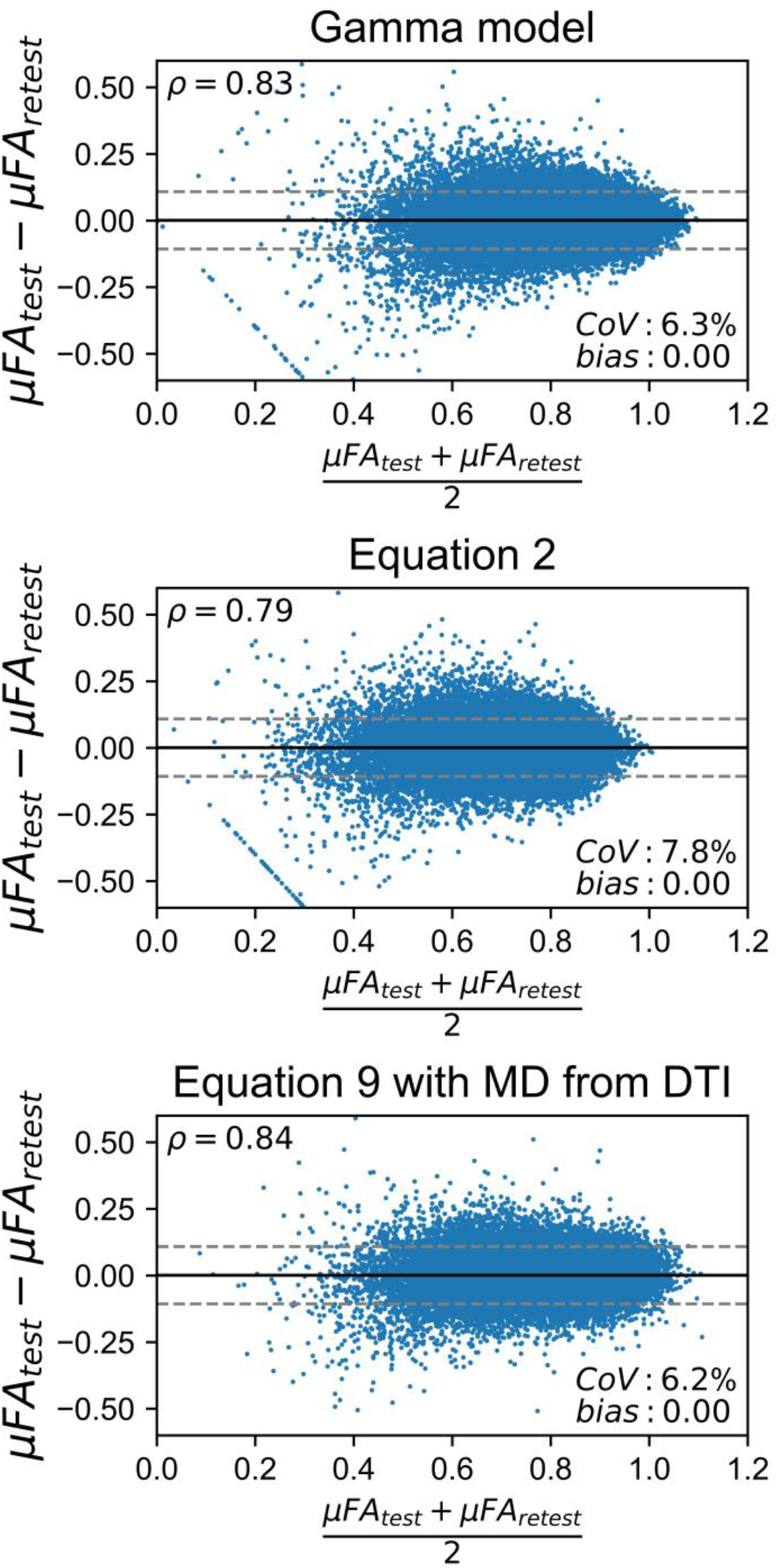
Bland-Altman plots assessing the test-retest reliability of μFA estimates acquired using different techniques. The solid black line represents the mean bias, and the dashed grey lines represent the ±1.96 standard deviation lines.

## 5. Discussion

Microscopic anisotropy mapping has been gaining popularity in neuroimaging studies because it provides a marker of tissue microstructure independent of orientation dispersion. The aims of this work were two-fold: (1) to develop a computationally-efficient method to estimate μFA and determine the optimal imaging parameters (b-value and *n*_*iso*_/*n*_*lin*_) needed to maximize image quality for a given scan time or number of acquisitions, and (2) to validate the techniques described in this work against the more established gamma model in a clinically-viable protocol. The first aim was achieved by directly estimating μA^2^ from the cumulant expansion of powder-averaged LTE and STE acquisitions and then estimating the SNR of μA^2^ using standard error propagation theory. The optimal b-value of 2000 s/mm^2^ falls within the optimal range for DDE methods; Ianus et al found that b-values between 2000 and 3000 s/mm^2^ are optimal for single-shell DDE estimations of μA because lower b-values result in noisy images while higher b-values result in large biases (Ianuş et al., 2018). The optimal *n*_*iso*_/*n*_*lin*_ (*S*_*lin*_*/S*_*iso*_) is somewhat intuitive as STE images typically have lower signal than LTE images due to the more rapid signal decrease with b-value. Notably, a steep drop-off in SNR with *n*_*iso*_/*n*_*lin*_ ratios below 1 was observed. These optimization findings were validated by the test-retest CoV ratio between the standard and suboptimal data sets agreeing with the SNR ratio predicted by equation (11). The second aim was achieved by acquiring all the data necessary for all the different μFA volumes in a single acquisition, mapping μFA from different subsets of data, and performing voxelwise comparisons on the maps. Notably, the direct approaches using equations (3) and (9) yielded comparable reliability and strong correspondence with the gamma method.

The μFA imaging techniques proposed in this work are suitable for clinical use due to the relatively minimalistic acquisition protocols needed to estimate μA^2^ and μFA. Using equation (2) or (9), μFA, isotropic kurtosis, and anisotropic kurtosis can be estimated from as few as 4 powder-averaged signals, with at least one acquisition being an STE acquisition and another being an LTE acquisition. Furthermore, μFA computation time in the standard subset only took approximately 2 minutes using the regression approach in the standard subset, versus approximately 120 minutes when using the gamma model, evidence. To demonstrate an even faster protocol than the standard protocol used for comparisons in this work, a further reduced subset of the data (16 LTE directions at b = 2000 s/mm^2^ and 3, 3, and 18 STE acquisitions at b = 100, 1000, and 2000 s/mm^2^, respectively) was used to estimate μFA, μA^2^, and the linear kurtosis using a joint non-linear least squares fit, shown in Fig. 8. Note that post-processing was performed on this subset after separating it from the rest of the data. This protocol would require only 3.3 mins of scan time and demonstrates that the linear kurtosis can be estimated from a set of data containing only one LTE shell when *D* is assumed to be the same between LTE and STE acquisitions.

**Fig. 8.**
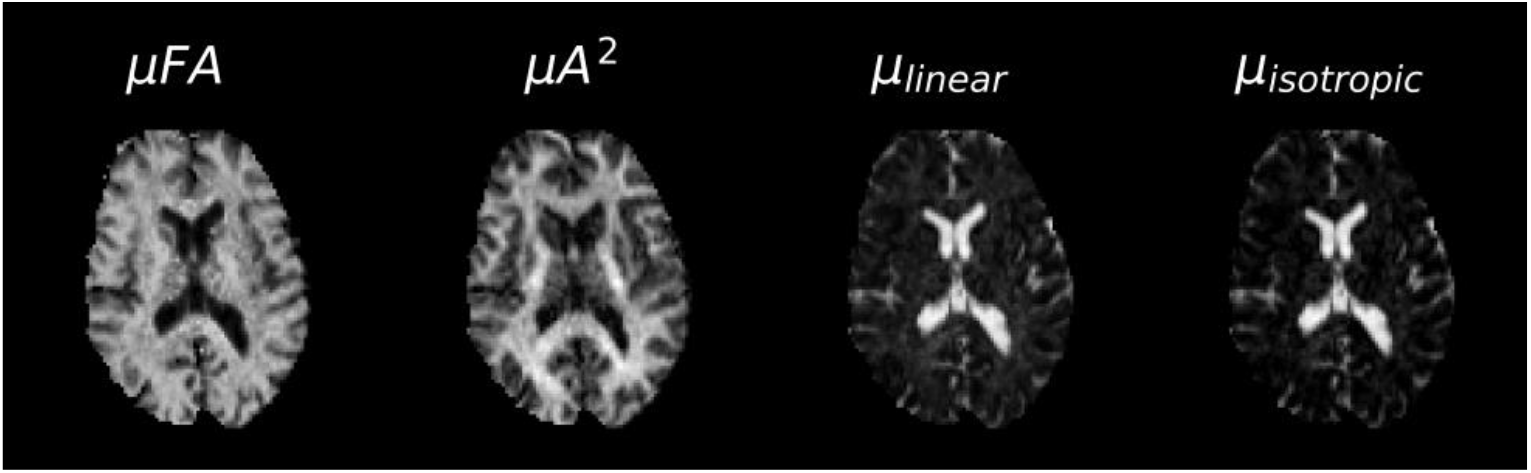
Example μFA, μA^2^, and linear and isotropic variance maps acquired using equation 2 in a subsampled data set: The acquisition comprised of 16 LTE directions at b = 2000 s/mm^2^ and 3, 3, and 18 STE directions at b = 100, 1000, and 2000 s/mm^2^, respectively. This direction scheme corresponds to a total scan time of approximately 3.3 min with 220 mm x 220 mm x 96 mm coverage at an isotropic 2 mm resolution. All images were normalized to a maximum pixel value of 1.

In this study, biases were observed in the μFA WM maps relative to the measurements produced by the gamma model. The μFA_E2_ metric had a large mean bias of −0.11 compared to μFA_gamma_, while the μFA_E9+DTI_ metric was biased against μFA_gamma_ by a modest −0.02. We suspect that the most likely causes of this discrepancy between the techniques are the differences between the models used to fit the data: the implementation of the gamma model used in this work utilizes a soft Heaviside function to constrain the fit to more heavily use the lower b-values, similar to the DTI fit for *D* in μFA_E9+DTI_. Accordingly, strong correspondence was observed between μFA_gamma_ and μFA_E9+DTI_. Using a full kurtosis fit to estimate D resulted in lower μFA values in the μFA_E2_ volume, which reveals a potential bias in the other two methods that results in physically implausible μFA values that are greater than 1 (see Fig. 7). That said, μFA computed from the equation (2) approach could also be biased to lower values because the cumulant expansions of the powder-averaged signals were limited to the second order (equations (4) and (5)), ignoring the effects of higher order terms. A previous study that used DDE to estimate μA at a single b-value in six different microstructural models (Ianuş et al., 2018) reported an underestimation of the metric when acquired at a single b-shell; to remove this bias, the use of a multiple b-shell approach to remove higher order cumulant terms can be considered.

Qualitatively, the biases between the different volumes did not have a significant impact on the images as contrast between structures or regions and image quality appeared similar in all the maps. Additionally, voxel-wise comparisons between the maps showed strong linear relationships in WM regions, evidence that the biases between the different techniques are likely scalar or constant. We propose that each of the techniques described in this work may be suitable for use in clinic or research under the caveat that studies assessing multiple patients or assessing patients longitudinally should use the same protocol and technique to avoid biases.

There are several limitations potentially affecting the accuracy of this study. The STE sequence used in this work utilizes different gradient waveforms in each diffusion-encoding direction, probing each at slightly different diffusion times and over different trajectories in q-space. Given the small microstructural length scales in WM (<10 μm), the long diffusion time regime is likely an appropriate assumption for all 3 waveforms. However, it may be worthwhile to investigate the individual waveforms separately in phantoms or in vivo acquisitions, or to swap the gradients (e.g. Gx with Gy) to test for inconsistencies in resulting images.

Another limitation is the protocol optimization cohort of volunteers, which contained 4 people, all under the age of 25. Water diffusion anisotropy has been shown to decrease with aging (Hsu et al., 2010; Sullivan and Pfefferbaum, 2006) and thus this cohort represents a group in which μA and μFA in brain tissue could be expected to be higher, on average, than in older subjects. Nevertheless, the resulting optimal b-value of 2000 s/mm^2^ is consistent with the values used in previous studies that used single-shell DDE acquisitions and those that used multi-shell nonconventional protocols.

A relatively low number of LTE directions were acquired at b = 2000 s/mm^2^ in the optimized noisy and standard denoised subsets, which may have slightly reduced the accuracy of the measurements by introducing a directional dependence to the powder-averaged signal <u>(Nilsson et al., 2020)</u>. This would not have affected comparisons between μFA_E2_ and μFA_gamma_, but the μFA_E9+DTI_ volume was computed with more STE acquisitions at b = 2000 s/mm^2^, which may have slightly advantaged measurements from that volume against the others, particularly in the test-retest analysis.

## 6. Conclusion

In conclusion, we have demonstrated an optimized model-free technique that enabled full-brain mapping of μA and μFA in a clinically relevant 3.3 min scan time at 3T. Two implementations of the proposed direct approach were validated against the gamma model, and an approach to determine the optimal maximum b-value and ratio of STE to LTE acquisitions was proposed and validated. Compared to other μFA techniques involving the use of nonconventional pulse sequences, the direct method requires fewer b-shells (and, thus, fewer total directions) and significantly less computation. Though additional work is necessary to establish the roles of μA and μFA imaging in clinical settings, the ability to rapidly probe these measurements in vivo opens the door for exploration into their abilities to assess neurodegeneration and other pathologies.

## Data Availability

Data will be made available and should be referenced as follows: Baron, C., Arezza, N.J.J., 2020. Test-Retest Data Repository for Spherical Tensor Encoding [WWW Document]. URL https://osf.io/etkgx/

## Appendix Signal to noise ratio of μA^2^ estimation

The variance of μA^2^ (*σ*^*2*^_*μA2*_), assuming equal noise in STE and LTE images and that there is no covariance between the two acquisition types, can be approximated using the error propagation equation. Propagating error from equation (8) yields:

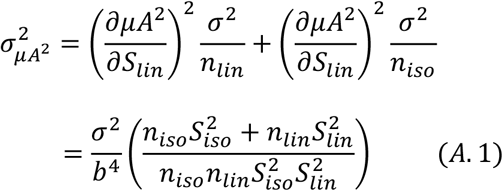

where *σ* is the noise in an STE or LTE diffusion-weighted MR image, *b* is the b-value, *n*_*lin*_ is the number of LTE directions acquired, *n*_*iso*_ is the number of STE averages acquired, and *S*_*lin*_ and *S*_*iso*_ are the mean signals in LTE and STE acquisitions, respectively. The SNR of a μA^2^ image or volume (*SNR*_*μA2*_) can be estimated as the μA^2^ metric divided by its standard deviation:

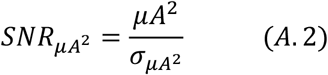

Substituting equations (8) and (A.1) into (A.2) yields equation (11):

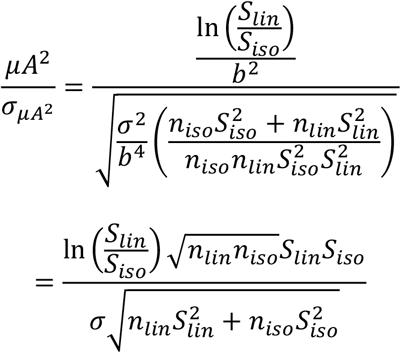

To determine the optimal ratio of *n*_*iso*_*/n*_*lin*_ as a function of the mean LTE and STE signal at a single b-value, we can express equation (A3) in terms of only *n*_*iso*_ and *n*_*lin*_, replacing most other terms with the constant *C*. We can also confine the total number of acquisitions to an integer value, *T*, and replace *n*_*iso*_ with *T-n*_*lin*_ to reduce the number of unknown variables in the formula. The resulting expression is:

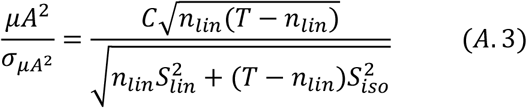

The maxima and minima of equation (A.3) can be calculated by solving for the roots of the derivative of the SNR equation:

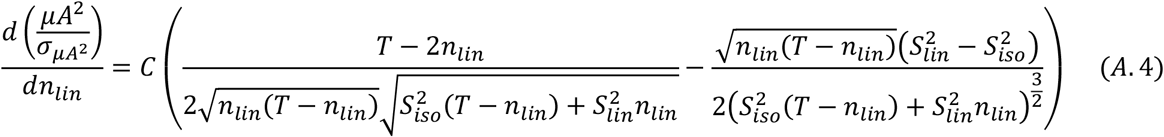

The roots of (A.4) are *n*_*lin*_ *= TS*_*iso*_*/(S*_*iso*_ *-S*_*lin*_) and *n*_*lin*_ *= TS*_*iso*_*/(S*_*iso*_ *+ S*_*lin*_), the prior of which is not realizable because *n*_*lin*_ would be negative if *S*_*iso*_ *< S*_*lin*_. Rearranging the latter yields the optimal ratio of STE to LTE acquisitions:

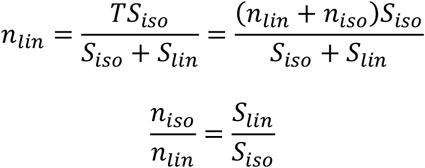

## Acknowledgements

This work was supported by the Natural Sciences and Engineering Council of Canada [grant number RGPIN-2018-05448], Canada Research Chairs [number 950-231993], Canada First Research Excellence Fund to BrainsCAN, and Ontario Graduate Scholarships. The authors would also like to thank Ms. Naila Rahman (Western University) for aiding with image registration.

